# Immune response after SARS-CoV-2 infection with residual post COVID symptoms

**DOI:** 10.1101/2022.10.10.22280762

**Authors:** Tanyaporn Pongkunakorn, Thamonwan Manosan, Apinya Surawit, Suphawan Ophakas, Pichanun Mongkolsucharitkul, Sureeporn Pumiem, Sophida Suta, Bonggochpass Pinsawas, Nitat Sookrung, Nawannaporn Saelim, Kodchakorn Mahasongkram, Pannathee Prangtaworn, Anchalee Tungtrongchitr, Watip Tangjittipokin, Kobporn Boonnak, Tassanee Narkdontri, Nipaporn Teerawattanapong, Anan Jongkaewwattana, Korapat Mayurasakorn

**Affiliations:** Siriraj Population Health and Nutrition Research Group, Department of Research and Development, Siriraj Medical Research Center, Mahidol University, National Science and Technology Development Agency, Pathumthani, Thailand; Center of Research Excellence on Therapeutic Proteins and Antibody Engineering, Department of Parasitology, Faculty of Medicine Siriraj Hospital, Mahidol University, National Science and Technology Development Agency, Pathumthani, Thailand; Department of Immunology, the Faculty of Medicine Siriraj Hospital, Mahidol University, National Science and Technology Development Agency, Pathumthani, Thailand; Research Department, Faculty of Medicine Siriraj Hospital, Mahidol University, National Science and Technology Development Agency, Pathumthani, Thailand; National Center for Genetic Engineering and Biotechnology, National Science and Technology Development Agency, Pathumthani, Thailand

## Abstract

**BACKGROUND:** In a number of patients, post-acute COVID syndrome develops after acute infection with severe acute respiratory syndrome coronavirus 2 (SARS-CoV-2) (Long COVID [LC]). Here, we examined the immune responses and clinical characteristics of individuals with LC compared to age- and gender-matched healthy recovered COVID individuals (HC) during the Omicron pandemic. Immune responses following BNT162b2 (Pfizer) booster are also determined.

**METHODS:** This retrospective cohort study included 292 patients (LC, 158; HC, 134) confirmed to have SARS-CoV-2 infection from January to August 2022. We determined anti-SARS-CoV-2 receptor-binding domain immunoglobulin G (anti-RBD IgG), surrogate virus neutralization test (sVNT), T-cell subsets, and neutralization of wild-type, BA.1 and BA.5. A subset of patients was voluntarily recruited for booster vaccination with BNT162b2 vaccine and immunogenicity was assessed 4weeks after vaccination.

**RESULTS:** Cycle thresholds were higher in the HC group than in the LC group (20.7 vs. 19.7; P<0.039). Anti-RBD IgG was higher at ≤56 days after COVID-19 onset (PC) in 3-dose vaccines compared with 2-dose vaccines in the LC group (P=0.02) and after 57-84 days PC in 3-dose vaccines in the HC group (P<0.001). The sVNT in LC was significantly high against Wuhan and sVNT was 30% lower against the Omicron than the Wuhan. sVNT was relatively sustained in 3-dose vaccines than ≤ 2-dose vaccines. sVNT in the HC group reached its peak at 57-84 days PC as compared with the LC group.

**CONCLUSIONS:** These findings imply that LC produced increased neutralizing antibody responses than those with HC. During the Omicron pandemic, immunity after LC has still waned; therefore, a booster vaccine may be needed after 2-3 months from last infection. (ClinicalTrials.gov number, NCT05484700)

## INTRODUCTION

Severe acute respiratory syndrome coronavirus 2 (SARS-CoV-2) is constantly changing and accumulating mutations in its genetic code over time.^1^ Omicron lineage (B.1.1.529) which has been acknowledged in November 2021 as a variant of concern (VOC), unprecedentedly spread to become globally dominant and later has evolved into two more transmissible BA.5 and BA.4 sublineages.^2^ BA.5 and BA.4 share many genetic mutations with BA.2.^2^ Omicron displaced Delta as the predominant variant during the study period. Randomly selected SARS-CoV-2 variants captured by surveillance conducted by the Department of Medical Science, Thailand, the UK Health Security Agency (UKHSA) and Centers for Disease Control and Prevention, USA during weeks 37 to 38 of 2022 demonstrated that almost all new infections in Thailand^3^, the U.K.^4^ and the U.S.^5^ were due to Omicron BA.5/BA.4 (49.2% - 95.5% - 96.4%) and Omicron BA.2 (59.6% - 3.4% - 1.4%). Now BA.2.75 was designated as a new variant of interest and is monitored. It would be investigated once there is a significant public health impact.^4^

Variants’ transmission including BA.5 and BA.4 which carry their own unique mutations can lead into a wide range of clinical presentation and biological modifications. These speed infectivity to infect more people even in fully vaccinated, boosted individuals as well as who were immune to earlier forms of Omicron and other variants.^1^ Most individuals with breakthrough SARS-CoV-2 infection can experience asymptomatic or mild-to-moderate-to-severe coronavirus disease 2019 (COVID-19).^6,7^ A previous study^1^ showed sera from BA.1 vaccine breakthrough infection exhibited significant reductions in neutralization against BA.5/BA.4; thereby, raising the possibility of Omicron reinfections.

Several lines of evidence show that there are multisystemic humoral responses and immune responses during acute illness and following recovery from the acute phase of COVID-19 infection. These contribute to both host defense and pathogenesis of severe COVID-19 and residual post COVID syndrome (Long COVID [LC]). Distinct immune dysregulation with lymphocytopenia, increased neutrophils and activation of the coagulation cascade have been described in the acute phase.^6,8^ Following acute COVID-19 infection, 10-70% of COVID-19 patients reported physical and mental symptoms 2-3 months after infection.^9-11^ Numbers of hospitalized symptomatic COVID-19 patients experienced LC symptoms. Still, more than half of patients with prior mild-to-moderate COVID-19 have LC persisting more than 4 months after infection.^12,13^ LC manifests as diverse symptoms affecting various organs-as seen in COVID-19 patients who did not recover fully.^14^ The most predominant LC symptoms are headache, breathlessness, cough, chest pain, abdominal symptoms, myalgia, fatigue, cognitive difficulties, anxiety and depression^15,16^ and neurologic/psychiatric symptoms.^17^ Evidence demonstrates that T cells and sustained immune dysfunction following even mild COVID-19 be associated with LC.^18^ For instance, effector molecule expression in memory T cells was reduced in neurologic patients with LC symptoms and sustained increases in T cell activities to SARS-CoV-2 mRNA vaccination was observed in this group compared with healthy COVID convalescents (HC).^19^ Nevertheless, the driver of this immune dysfunction and the immune response after vaccination, specific effects on LC are needed for further investigation and verification.

## RESULTS

### DEMOGRAPHIC CHARACTERISTICS AND CLINICAL MANIFESTATION

292 participants following COVID-19 infection enrolled from the surrounding Bangkok area. This included 158 participants (86.1% women) in the LC group and 134 participants (66.4% women) in the HC group. The mean age of both groups was similar (Table 1). Among those in the HC and LC groups, 130 (97%) and 157 (99.4%) were treated in out-patient home isolation system indicating that COVID-19 symptoms were mild-to-moderate. The median peak viral RNA based on Ct values in the LC groups (19.7 [IQR=18.0-21.9]) was lower than ones in the HC groups (20.7 [IQR=18.2-25.1]; P<0.037). The median of a targeted COVID-19 gene for envelope (E) was 18.4 (IQR = 16.3-23.3) for HC group and 17.8 (IQR = 16.5-19.9) for LC group (P=0.017). Our results showed no significant correlation between vaccination status (either primary two doses or booster shots) between groups (ns). Retrospective analysis revealed that patients in the LC group had a higher proportion of Favipiravir treatment (65%) than in the HC group (53.7%). This finding indicates that having LC was associated with lower Ct values and disease severity based on the prevalence of Favipiravir treatment. In the LC group, the presence of comorbidity was found in LC group more than HC group including hypertension (P=0.052) and obesity (P=0.025). It was found that most participants received more than three doses of vaccines (93.0% in LC vs. 90.3% in HC, P=0.306). The major residual LC symptoms included fatigue/myalgia (91.8%), breathlessness (74.7%), anorexia (55.1%), and problem with concentration (77.8%) and memory (63.9%). Other respiratory, gastrointestinal, and musculoskeletal symptoms were more reported in the LC groups than in the HC group (P<0.05).

**Table 1.**
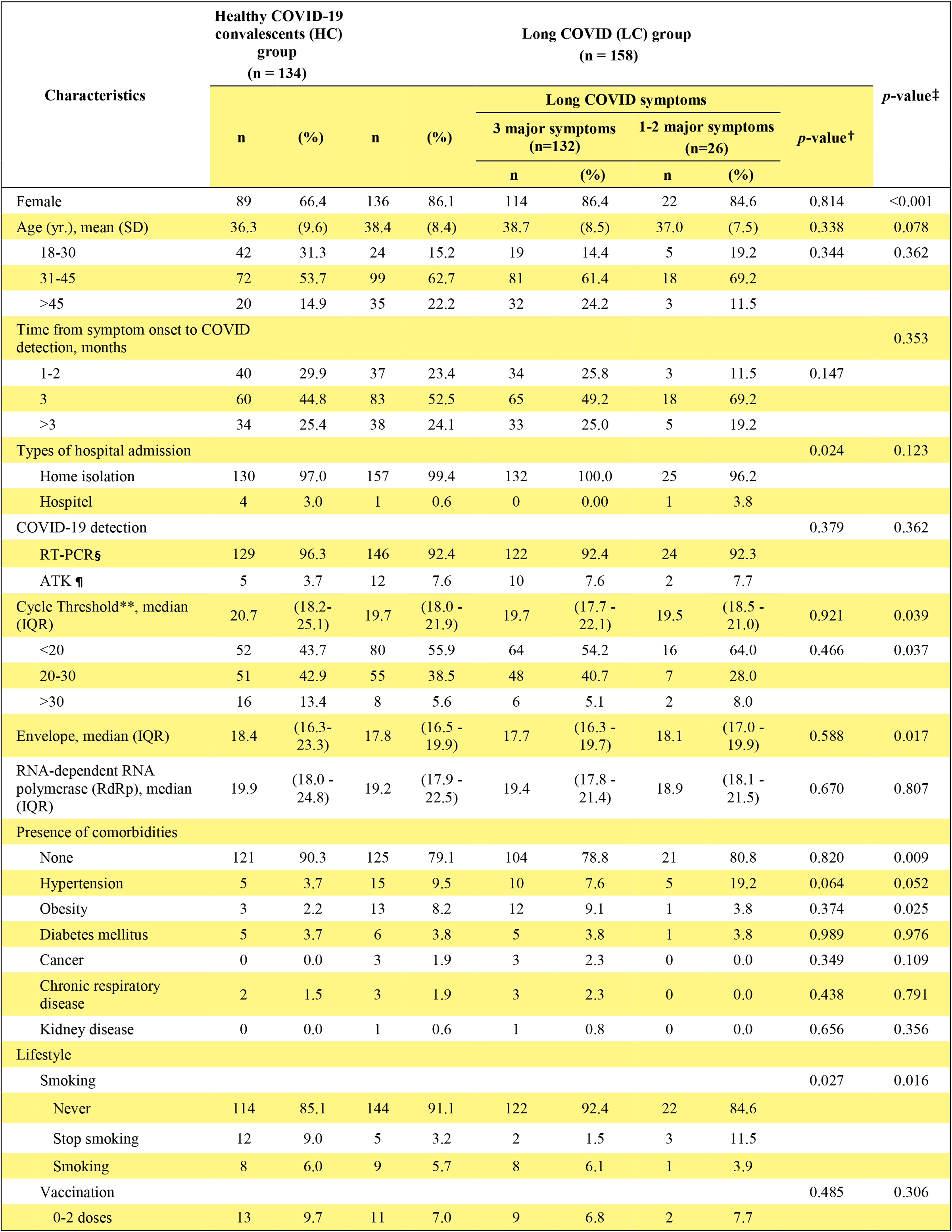
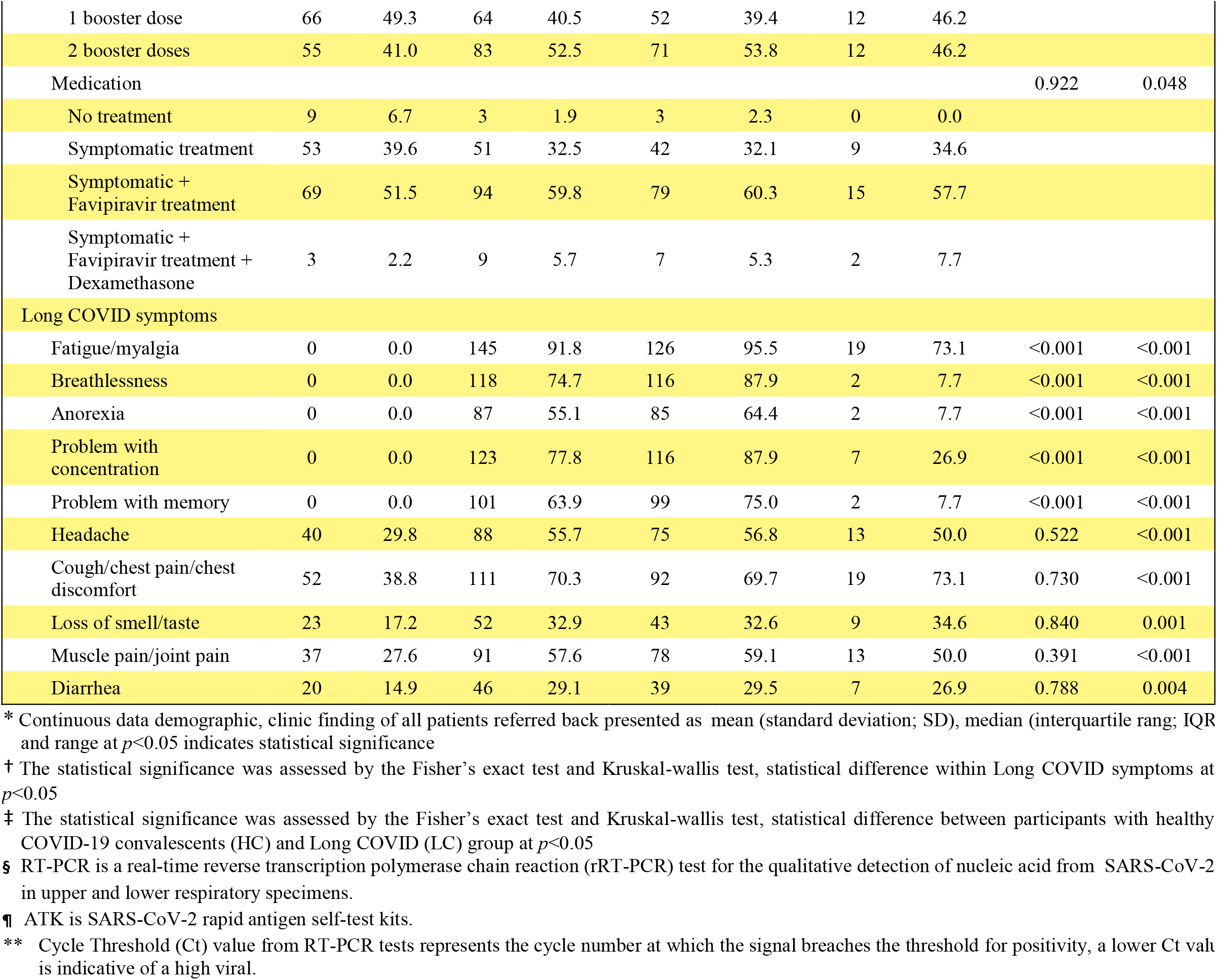
Demographic and clinical characteristics of participants*

### IMMUNE RESPONSES AGAINST SARS-COV-2 VARIANTS

High antibody titers were observed in both LC and HC groups (Figure 2A). The IgG antibody positive rates to SARS-CoV-2 S protein were found in nearly 100% at any time points in both LC and HC groups, except in one patient with 2 doses of vaccines prior to breakthrough COVID-19 infection who had negative IgG level at > 84 days. At least 3 doses of vaccines produced remarkedly high and comparable IgG levels between LC and HC groups (ns) suggesting that the titer of anti-SARS-CoV-2 IgG antibody was still detectable at high levels between two groups. Serum samples at different time points after illness onset showed differences in the distribution of IgG. The RBD-IgG geometric mean titers (GMT) were higher in 3-dose vaccines in the LC group at ≤ 56 days post COVID-19 infection (PC, P=0.02) as compared with 2-dose vaccines and in 3-dose vaccines in the HC group at 57-84 PC (P<0.001). The overall proportion of sera with an IgG > 1000 BAU/mL decreased from <56-day to 57-84-day and >84-day PC (86.8% to 79.6% and 74.7%, ns for HC vs. LC [Figure 2A]). This implied that recent booster vaccination enhanced IgG responses effectively and IgG antibodies were still remarkably high after three months PC. And the titers reached their peak around 2 months PC and decreased slightly (about 25%) over the following 3 months (GMT 2713 to 2085 in LC group and GMT 2482 to 2377 BAU/mL in HC group). Sera from individuals presumably recently infected with BA.5/BA.4 (Figure 2A [black dots]) showed high RBD-IgG GMT levels 4630 BAU/mL) as compared with other groups (Figure 2A [pink and blue dots], P<0.001). This implies that previous infection with BA.5/BA.4 but not an older Omicron variant (such as BA.1, BA.2) offers substantial more immune response regardless of individual vaccine heterogenous profile.

**Figure 1.**
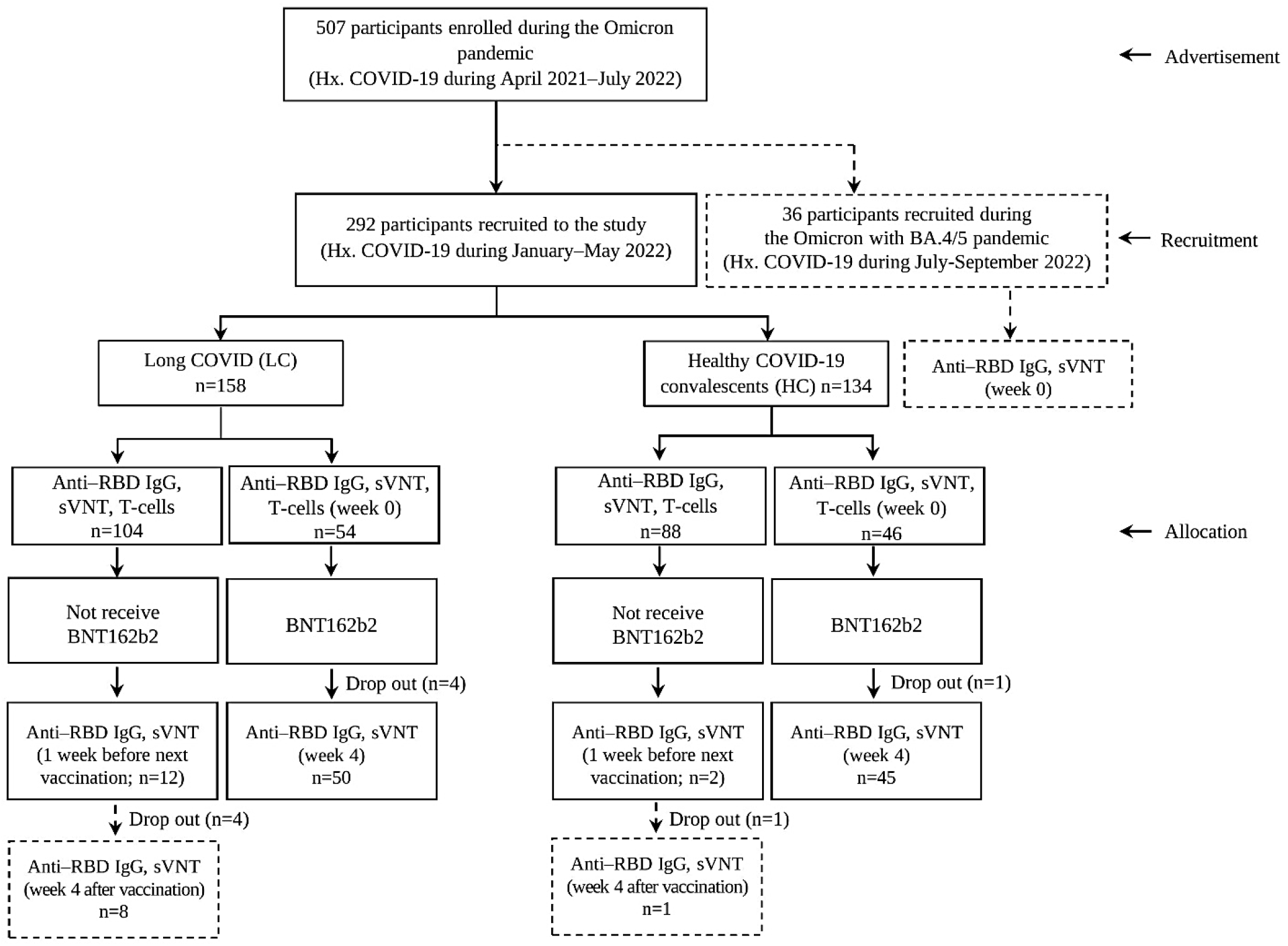
Study source recruitment and enrollment

**Figure 2.**
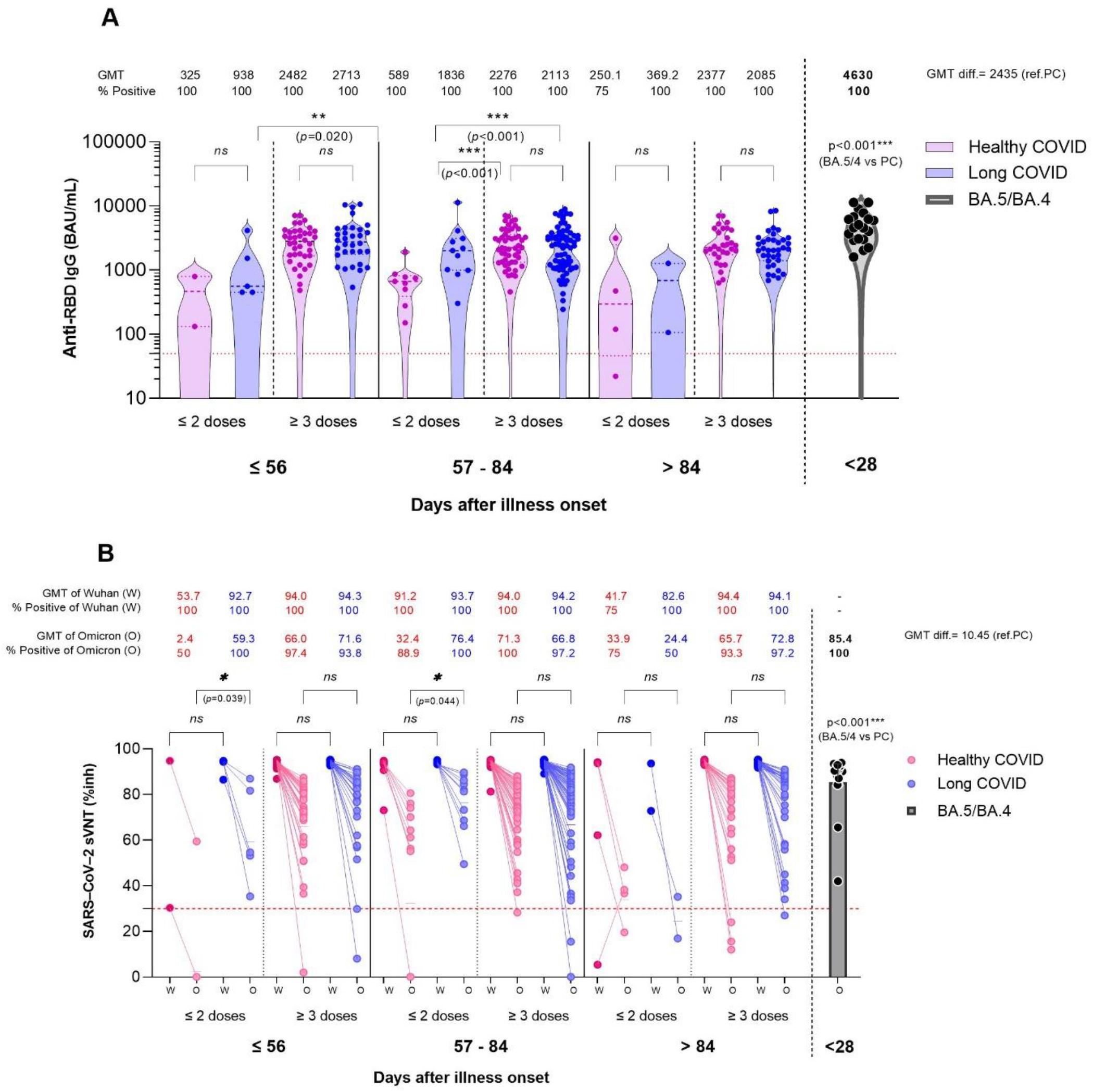
Immune responses after breakthrough COVID-19 infection with prior vaccination as compared between patients with healthy COVID-19 recovery (HC, pink dots), patients with residual long COVID (LC, blue dots) during the Omicron pandemic and patients during the BA.5/BA.4 (black color). (**A**) Geometric mean titers (GMTs) of SARS-CoV-2 anti-spike protein receptor-binding domain antibodies’ (Anti-RBD IgG) concentrations in serum samples obtained from subjects after COVID-19 infection and with prior various vaccination status and duration post COVID-19 onset (PC). All sera were from the patients during the Omicron pandemic. The dotted line (red) represents the threshold for positive in this assay. **(B**) Scatter plots demonstrate an inhibition rate of Wuhan and Omicron RBD-blocking antibodies measured using a surrogate viral neutralization test (sVNT) by vaccination/reinfection status; the lower dot line represents the cut-off level for seropositivity. The generalized linear model (GLM) was applied to evaluate the association between immune response (including antibody IgG and sVNT responses) and potential factors (i.e., vaccination, sex, and age). * P<0.05; ** P<0.01; **** P<0.0001.

### NEUTRALIZING ANTIBODY RESPONSES

Ninety nine percent of the patients had positive sVNT against wild type and Omicron (Figure 2B). In the LC group, sVNT was high against Wuhan regardless of doses of vaccines and at any time points PC even at >84-day PC, whereas positive sVNT titers were chiefly observed since at <56 days PC (Omicron GMT 59.3%) and sVNT was approximately 30% lower against the Omicron variant in most time points as compared with the Wuhan. sVNT was relatively sustained in 3-dose vaccination than ≤ 2-dose vaccination. In the HC group, sVNT was also high against Wuhan regardless of doses of vaccines and at all time points PC. However, sVNT against the Omicron was also 30% lower than those against Wuhan. Of note, sVNT in the HC group reached its peak at 57-84 days PC as compared with the LC group, sVNT of which was still at its high level after 84 days. This can imply that LC produced increased and more sustainable neutralizing antibody responses than those with full recovery. Additionally, those with recent BA.4/BA.5 breakthrough infection showed higher SVNT titers against the overall Omicron variant. This benefit has helped to prevention reinfection at least 2-3 months PC in the LC group and in those with BA.4/BA.5 prior infection.

### IMMUNE RESPONSE AFTER A VACCINE BOOSTER

The immunogenicity against SARS-CoV-2 after a booster dose of the Pfizer-BioNTech mRNA vaccine measured at 4 weeks after the vaccination in a subgroup of participants were analyzed. IgG antibodies directed against RBD were double after a booster vaccine (P<0.0001). However, the average GMT concentration of anti-RBD IgG in the LC group was 10% higher than that in the HC group (4665±1.76 vs. 4152±2.15 BAU/mL, P<0.05) (Figure 3A). Of note, sVNT against the Omicron variant was similarly increased in nearly all participants irrespective of having LC (LC 92.87% vs. HC 90.09%, ns [Figure 3B]). This implies that a booster mRNA vaccination may be beneficial to enhance immunity after COVID-19 infection especially for vulnerable individuals.

**Figure 3.**
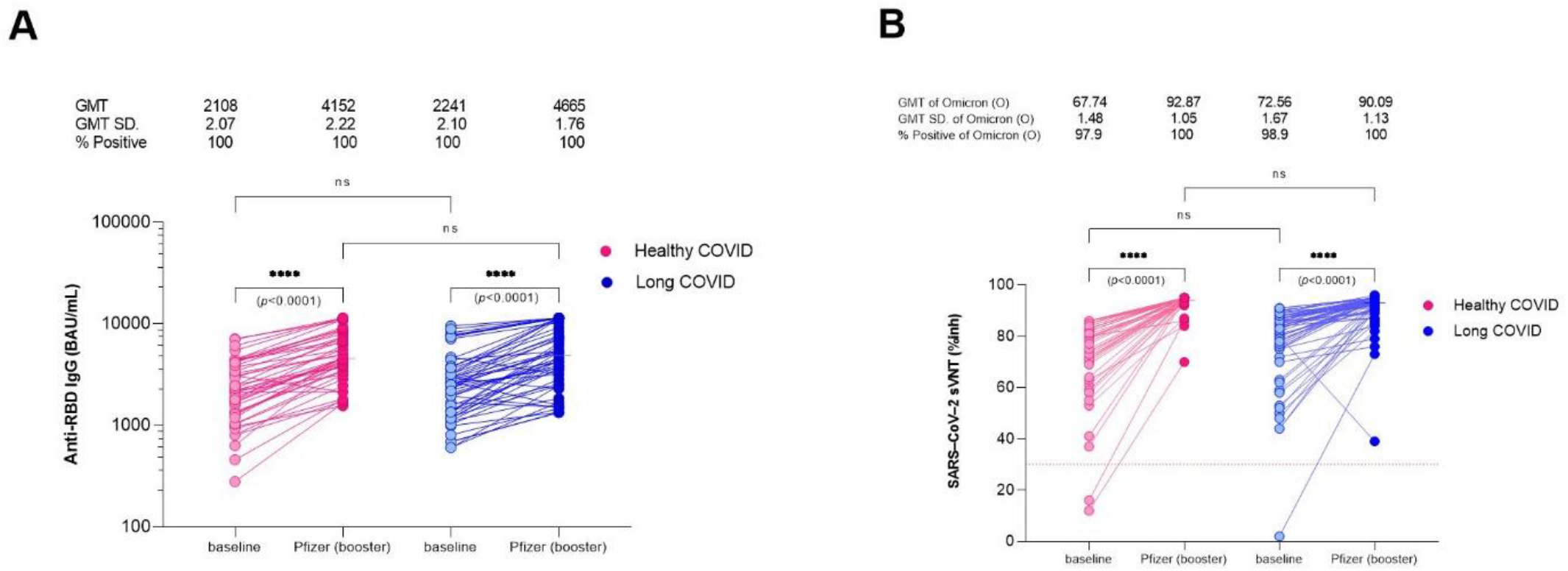
Immunogenicity against SARS-CoV-2 after a booster dose of the Pfizer-BioNTech mRNA vaccine compared between patients with healthy COVID-19 recovery (HC, red color) patients and patients with residual long COVID (LC, blue color) during the Omicron pandemic by scatter plots analysis. (**A**) A SARS-CoV-2 anti-spike protein receptor-binding domain antibodies’ (Anti-RBD IgG) concentrations in serum samples obtained from subjects after a 30-ug Pfizer booster vaccine at 4 weeks after vaccination. **(B)** An inhibition rate of the Omicron RBD-blocking antibodies measured using a surrogate viral neutralization test (sVNT). The generalized linear model (GLM) was applied to evaluate the association between immunogenicity of anti-RBD IgG and sVNT, and the potential factors (i.e., sex and age). * P<0.05; ** P<0.01; **** P<0.0001.

## DISCUSSION

In this study, we ascertained the following central findings. First, LC presents as a constellation of enervating symptoms most commonly including unremitting fatigue/myalgia, post-exertional malaise, breathlessness, anorexia, and cognitive dysfuntion.^20^ In accordance with previous studies, this syndrome was more prevalent in women, middle-aged patients and in patients with even mild-to-moderate COVID-19 irrespective of vaccination status.^9,21^ Age is recognized as the general determinant of disease severity mainly because the immune response deteriorates in aging people.^22^ With regard to the uncertainty about the frequency and prevalence of such individual symptoms of LC and how long they last, several patients reported fatigue shortly after COVID-19 recovery. A systematic review showed that self-report fatigue after recovery from COVID-19 infection can last up to 6 months and immunological dysfunction can persist for 8 months post mild-to-moderate COVID-19.^18,20^ SARS-CoV-2 may remain within some patients with the long-term symptoms, thereby leading to chronic inflammation and some organ and tissue dysfunction.^23^ In a previous cohort study, chronic post-COVID “brain fog” had significant associations with sex (female), respiratory symptoms at the onset, and the severity of the illness.^24^ However, increasing evidence indicates that LC can develop regardless of the severity of the original symptoms^25^ but it may depend on antigen persistence and sustained specific immune responses to SARS-CoV-2.^26^

Second, a well-defined understanding of innate and adaptive immune response in LC in association with vaccination status and heterogeneous characteristics of the virus and host responses is indispensable for planning for therapeutic and vaccination strategies. Our results showed that there were no significant differences in immune response between LC and individuals with complete recovery, regardless of vaccination status before COVID-19 infection. Significant decreases in levels of IgG and neutralizing antibodies were also observed among participants without booster vaccines prior to COVID-19 infection, which are classically associated with waned immunity. High levels of antibodies were still remarkably observed after three-month PC. An analysis of immune responses of those who were presumably infected with BA.5/BA.4 displayed significant increases in the immunity as compared with those who were infected with other Omicron variants. More interestingly among a subset of participants with LC, anti-RBD IgG antibody but not sVNT after a Pfizer-BioNTech booster shot was significantly increased compared with the HC group. These findings are supported by prior reports that LC pathogenesis might include persistent viral antigen, reactivation of latent herpesviruses, and chronic inflammation, which may be associated with the elevation in antibody responses.^11,27^

There were some limitations in this study. During the study period, most COVID-19 cases had mild-to-moderate symptoms and they were most likely to self-quarantine at home. Therefore, we only had patients under home isolation conditions, and it does not provide a fair comparison about the long COVID in the hospitalized patients and the results could not be generalized to all types of long COVID community. In addition, our study focused on post-acute residual COVID-19 symptoms at 1-4 months post. The onset of COVID-19; therefore, it cannot discern whoever will develop long-term chronic LC. An additional limitation is sample size. Patients who exhibited both a classic LC symptom and risk factors may constitute only a minute subset, making it difficult to draw specific, novel predictors.

## CONCLUSION

No clinical and laboratory differences have been identified between participants with long COVID and demographically and medically matched convalescent and healthy control groups and all were responded remarkedly after a booster dose of an mRNA vaccine after the COVID-19 infection. Sera from participants infected during the Omicron BA.5/BA.4 but not the Omicron BA.1, BA.2 showed the better immune responses against the overall Omicron variants. Our study provides a basis of immunological and evidence for vaccination program after COVID-19 infection with or without long COVID symptoms.

## METHODS

This retrospective cohort study compared immunogenicity post COVID-19 between individuals with residual LC symptoms and ones with HC related to pathophysiology, immunology, and clinical consequence. In addition, immune responses at 4 weeks were determined after a subset of these participants was voluntarily administered with additional 30 ug of BNT162b2 (Pfizer) (Figure 1). The Institutional Review Board (IRB), Faculty of Medicine Siriraj Hospital, Mahidol University reviewed and approved the follow-up study (COA no. Si 1036/2021).

### STUDY POPULATION

All patients (age 18-65 years) were invited for volunteer participation in the study of the immune responses against SARS-CoV-2 after breakthrough infection. The eligible participants were those who had confirmed SARS-CoV-2 infection based on a patient’s medical certificate from January to May 2022 (during the Omicron-dominant pandemic), and they had not received any additional COVID -19 vaccine after infection. Written informed consent was obtained from all participants. Pregnant participants and people having a history of allergic reactions after COVID-19 vaccination, or any drugs were excluded. Participants were defined as having LC when they had residual self-reported LC symptoms longer than 1-month after COVID-19 onset. LC symptoms were based on the presence of one of the three major symptoms: fatigue/myalgia, breathlessness, and anorexia.^18^ On the other hand, patients without LC symptoms were called “healthy COVID-19 recovery; HC”. These two groups were gender and age (±5 years) matched with each other (Table 1). Based on their reported residual symptoms from questionnaires, 158 participants with LC symptoms and 134 participants with HC were enrolled into the study. We performed a subset of analyses on an independent group of 36 post-acute COVID-19 patients (ambulatory adults infected during the Omicron with BA.5/BA.4 pandemic) to compare the immune responses with our primary cohort. Previous infections were classified as Omicron BA.1/BA.2 dominant versus Omicron BA.5/BA.4 dominant previous infections based on if they occurred before or after the emergence of BA.5/BA.4 wave that started in Thailand since June 2022.^3^

### A BOOSTER AND IMMUNOGENICITY

In the studied cohort, a subset of 100 patients (age≥18 years) were voluntarily recruited for immunogenicity follow-up study after receiving 30 ug of BNT162b2 (Pfizer) as a booster shot at 29-84 days post COVID-19 onset. All participants provided informed consent for this study. They were tested for SARS-CoV-2 antibodies and a surrogate virus neutralization (sVNT) against SARS-CoV-2 Wuhan and Omicron variants. Those who denied this booster dose were asked to come back for IgG testing again before their next booster shot to determine levels of waned immunity. The Department of Disease Control, Ministry of Public Health (Thailand) kindly provided the Pfizer-BioNTech (Comirnaty) BNT162b2 vaccine, designated Lot. 1L085A. This RNA vaccines belong to a state-of-art approach that uses genetically engineered mRNA of viral spike proteins included in a lipid nanoparticle, which subsequently inject intramuscularly into human body and allow the ribosomes in the cell to synthesize the viral spike proteins that safely prompts an immune response.^28^ We further explore the immunogenicity of this boosting at 4-week post booster shot.

### OMICRON BA.5/BA.4 IMMUNOGENICITY

For the pilot study, 36 patients who were diagnosed with COVID-19 during July 15 to September 14, 2022, were recruited. This provided a proxy for BA.5/BA.4 infections, considering the incidence of this variants during the recruitment time. Immune responses against the overall Omicron variants were evaluated for viral neutralization against BA.5/BA.4.^3^

### OUTCOME MEASURES

The primary outcome was to compare patients’ baseline clinical and biological characteristics between LC and HC groups. The outcome included demographic data, clinical and biochemical characteristics. Participants shared a copy of their government-issued documents with study staff to verify their vaccination information. The “date of disease onset” was defined as the day when new-onset self-reported respiratory symptoms were observed. Viral loads were considered in cycle threshold (Ct) value analyses. Analyses considered viral load for comparisons of Ct values by vaccine exposure groups and self-reported symptoms as previously described.^6^

### SARS-COV-2 RT-QUANTITATIVE PCR ASSAY (RT-PCR)

Diagnosis of COVID-19 was made based on the detection of ≥2 SARS-CoV-2 genes by RT-PCR from nasopharyngeal (NP) swab, throat swab, and/or any respiratory samples as previously described.^29^ COVID-19 diagnostic assay was a probe-based qualitative RT-PCR probe. Allplex™ 2019-nCoV Assay (Seegene, Seoul, South Korea) was used for SARS-CoV2 detection. The targeted COVID-19 genes detected here included nucleocapsid (N), envelope (E) of Sarbecovirus and RNA-dependent RNA polymerase (RdRp) of COVID-19 according to the manufacturer’s instructions and described previously.^30^ Antigen test kit (ATK) was also accepted as a confirmation of COVID-19; in case that RT-PCR could not be assessed.

### SEROLOGICAL ASSAYS

Blood samples were collected at baseline (pre-booster), and 4 weeks after a booster shot (post-boost) in a subset of participants from both groups. Briefly, plasma samples were processed for anti-SARS-CoV-2 receptor-binding domain immunoglobulin G (anti-RBD IgG, (S1 subunit, No. 06S600), SARS-CoV-2 IgG II Quant for use with ARCHITECT; Abbott Laboratories, USA) as described previously.^6^ This assay linearly measures the antibody level between 21.0 - 80,000.0 arbitrary unit (AU)/mL, which was converted later to WHO International Standard concentration as binding antibody unit per mL (BAU/mL) following the equation provided by the manufacturer (BAU/mL = 0.142 x AU/mL). The level greater or equal to the cutoff value of 50 AU/mL or 7.1 BAU/mL was defined as seropositive.

A surrogate virus neutralization test (sVNT) was undertaken against the or wild type (Wuhan) strain and the Omicron (B.1.1.529) strain. Briefly, plasma was pre-incubated with horseradish-peroxidase-conjugated receptor-binding domain protein (HRP-conjugated RBD protein). Subsequently, the mixture was transferred to each well containing Streptavidin bound with Biotin-conjugated angiotensin-converting enzyme 2 (ACE2). Finally, the optical density absorbance was measured using a spectrophotometer at 450 nm. Sample diluent was used as the negative control. The inhibition rate was calculated through this formula:

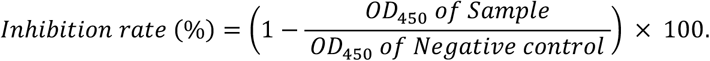

### PERIPHERAL BLOOD MONONUCLEAR CELLS ISOLATION AND COLLECTION

Isolation of peripheral blood mononuclear cells (PBMC) was done from peripheral blood supplemented with anticoagulants by using Ethylenediamin tetra-acetic acid (EDTA) tube.^31^ Briefly, blood was diluted 1:1 with phosphate buffer saline (PBS [HyClone™, Utah, USA]) pH 7.2 3 ml of a density gradient medium (Lymphoprep [STEMCELL Technologies, Germany]) were added into a 15 ml tube. After that, 10 of diluted blood samples were carefully layered on top and centrifuged at 400xg for 30 min at 25°C (Allegra X-15R Centrifuge, Beckman Coulter). The PBMC were transferred to a new 15 ml tube, and 10 ml of PBS were filled, mixed, and centrifuged at 400xg for 5 min at 25°C. The supernatant was completely removed, and the cells were resuspended with a small volume of PBS. When most of the platelets were removed, the cells were suspended in a complete cryoprotective media with 10% dimethyl sulfoxide (DMSO [SIGMA Life Science, USA]) and fetal bovine serum (FBS [Gibco™, Thermo Fisher Scientific Inc., USA]) in the cryovials. Finally, the cryovials were frozen at -80°C for 1 day and then were transferred and stored in liquid nitrogen.

### MICRONEUTRALIZATION ASSAYS USING THE AUTHENTIC SARS-COV-2 VIRUSES

Sera from participants were used to assess the neutralization against wild type (WA1), and Omicron (B.1.1.529, BA.2 and BA.5). SARS-CoV-2 isolates (kindly provided by Prof. Florian Krammer, Icahn school of medicine at Mount Sinai, NY, USA). All procedures were performed in a certified biosafety level 3 (BSL-3) facility following standard safety guidelines. In brief, Vero.E6-TMPRSS2 cells were seeded in 96-well high binding cell culture plates (Costar, cat. no. 07620009) at a density of 20,000 cells per well in complete Dulbecco’s modified Eagle medium (cDMEM) one day before the infection. Heat inactivated serum samples (56 °C for 1 h) were serially diluted (three-fold) in minimum essential media (Gibco, life technology, Grand Island, NY, USA) supplemented with 2 mM L-glutamine (Gibco, life technology, Grand Island, NY, USA), antibiotic-antimycotic (Invitrogen-Gibco, Carlsbad, CA, USA) and 0.2% fetal bovine serum (Gibco, life technology, NY, USA). Serially diluted sera were incubated with 10,000 TCID of SARS-CoV-2 viruses for one hour at room temperature, followed by the transfer of 120 μL of the virus–serum mix to Vero.E6-TMPRSS2 plates. Infection proceeded for 1 h at 37°C and inoculum was removed. One hundred μL per well of the corresponding antibody dilutions plus 100 μL per well of infection media supplemented with 2% fetal bovine serum (Gibco, life technology, NY, USA) were added to the cells. Plates were incubated for 48 h at 37°C followed by fixation overnight at 4°C in 200 μL per well of a 10% formaldehyde solution. For staining of the nucleoprotein, formaldehyde solution was removed, and cells were washed with and permeabilized by adding 150 μL per well of PBS with 0.1% Triton X-100 (Fisher Bioreagents, Pennsylvania, USA) for 15 min at room temperature. Permeabilization solution was removed, plates were washed with 200 μL per well of PBS (Gibco, life technology, NY, USA) twice and blocked with PBS, 3% BSA for 1 h at room temperature. Blocking solution was removed and 100 μL per well of biotinylated monoclonal antibody 1C7C7 (Center for Therapeutic Antibody Development at The Icahn School of Medicine at Mount Sinai ISMMS) at a concentration of 1μg /ml in PBS, 1% BSA was added for 1 h at room temperature. Cells were washed with 200 μL per well of PBS twice and 100 μL per well of HRP-conjugated streptavidin (Thermo Fisher Scientific, Massachusetts, USA) diluted in PBS, 1% BSA were added at a 1:2000 dilution for 1 h at room temperature. Cells were washed twice with PBS, and 100 μL per well of O-phenylenediamine dihydrochloride (Sigmafast OPD; Sigma-Aldrich, Massachusetts, USA) were added for 10 min at room temperature, followed by addition of 50 μL per well of a 3 M HCl solution (Thermo Fisher Scientific, Massachusetts, USA). Optical density (OD) was measured (490 nm) using a microplate reader. Analysis was performed using Prism 7 software (GraphPad). After subtraction of background and calculation of the percentage of neutralization with respect to the ‘virus-only’ control, a nonlinear regression curve fit analysis was performed to calculate the 50% inhibitory dilution (ID50), with top and bottom constraints set to 100% and 0% respectively. All samples were analyzed in a blinded manner.

## STATISTICAL ANALYSIS

Normally distributed continuous variables were summarized as the mean ± standard deviation (SD); otherwise, the median (interquartile range, IQR) was used. Categorical variables were described as percentages and compared using the Chi-squared test. Continuous variables were described using geometric mean antibody titers (GMT), median, and IQR. The Mann–Whitney U test was used to compare differences between groups. The generalized linear model (GLM) was applied to evaluate the association between immune responses (including anti-RBD IgG and sVNT) and potential factors (i.e., vaccination, sex, and age). The statistical significance of anti-RBD IgG, sVNT, and others was determined using Kruskal–Wallis and Dunn’s multiple comparisons tests using GraphPad Prism 9 (GraphPad Software, San Diego, CA, USA) and STATA version 17 (Stata Corp, College Station, TX, USA). Two-tailed P values less than 0.05 were considered significant (*P<0.05, **P<0.01, ***P<0.001 and ****P<0.0001).

## Data Availability

All data produced in the present study are available upon reasonable request to the authors

## ACKNOWLEDGEMENTS

The authors are grateful to the participants in this study. We would like to thank all the COVID-19 teams for their dedication and expertise in recruitment, specimen processing and biobanking. We thank Kantapat Boriuttham, Patcharaporn Thabsuwan, Pimchanok Boonariya, Thaksaphon Siripant, Supinya Piwlaor, Boonjira Wetprasit, Noppanat Wachiranan, Suavaluk Songlilitchuwong, and Sirapassorn Ditkasem from the Department of Biotechnology, Faculty of Applied Science, King Mongkut’s University of Technology North Bangkok for participant recruitment. We appreciate grant support from Faculty of Medicine, Siriraj Hospital, Mahidol University and the Department of Disease Control (DDC), the Department of Medical Services Ministry of Public Health (MoPH). Authors are very grateful for all support for this study: Tassanee Narkdontri and Nipaporn Teerawattanapong from Siriraj Center of Research Excellence for Diabetes and Obesity (SiCORE-DO); Utane Runpanich from the Department of Immunology; Population Health and Nutrition Research Group; Department of Parasitology; and Center of Research Excellence on Therapeutic Proteins and Antibody Engineering. We acknowledge Prof. Florian Krammer and Dr. Juan Manuel Carreno, Icahn School of Medicine at Mount Sinai, NY, USA for their contribution to microneutralization assay. We also would like to acknowledge Professor Kulkanya Chokephaibulkit, professor of Pediatric Infectious Diseases and Director of the Siriraj Institute of Clinical Research (SICRES) for her guidance for initiation of this project.

## AUTHORS CONTRIBUTIONS

T.P., T.M., S.O. and A.S. contributed to the experimental design, collected patient data, performed data experimentation and interpretation. T.P., K.M., and N.K. drafted and finalized the manuscript. A.S. performed the statistical analysis. W.T. performed the anti-RBD IgG testing and provided data establishing testing. Ni.S., Na.S. and A.T. performed sVNT testing. Ko.M. and P.P performed T cell immunity analysis. A.J. performed a pseudotyped virus neutralization test. K.B. performed microneutralization assays using live virus. K.B. and A.J. proofread. All authors read the manuscript and agreed with its content. The corresponding author (K.M.) attests that all listed authors meet authorship criteria and that no others have been omitted.

## DATA AVAILABILITY

Raw data used in this study, including de-identified patient metadata and test results, are available upon request.

